# A meta-analysis of Early Results to predict Vaccine efficacy against Omicron

**DOI:** 10.1101/2021.12.13.21267748

**Authors:** David S. Khoury, Megan Steain, James A. Triccas, Alex Sigal, Miles P. Davenport, Deborah Cromer

## Abstract

In the studies to date, the estimated fold-drop in neutralisation titre against Omicron ranges from 2- to over 20-fold depending on the study and serum tested. Collating data from the first four of these studies results in a combined estimate of the drop in neutralisation titre against Omicron of 9.7-fold (95% CI 5.5-17.1). We use our previously established model to predict that six months after primary immunisation with an mRNA vaccine, efficacy for Omicron is estimated to have waned to around 40% against symptomatic and 80% against severe disease. A booster dose with an existing mRNA vaccine (even though it targets the ancestral spike) has the potential to raise efficacy for Omicron to 86.2% (95% CI: 72.6-94) against symptomatic infection and 98.2% (95% CI: 90.2-99.7) against severe infection.

---

On 25^th^ November 2021 the SARS-CoV-2 variant Omicron (B.1.1.529) was declared a Variant of Concern (VOC) by the World Health Organisation. Omicron is proposed to escape immune recognition from existing vaccines due to the number of mutations in the spike protein, targeted by most of the widely available vaccines (1). We have previously developed a method for estimating vaccine efficacy based on neutralisation titres and shown that once the loss of neutralisation against a novel VOC is known it is possible to reliably predict vaccine efficacy against that variant (2, 3). On 7^th^ December 2021 the first data demonstrating a drop in neutralisation titres against Omicron emerged from the Sigal Lab (4) and was closely followed by similar data from Sheward et al. (5), the Ciesek Lab (6) and Pfizer Inc. / BioNTech SE (7, 8). Here, we use these early reports to predict vaccine efficacy against Omicron using our previously established approach (2, 3).

In the studies to date, the estimated fold-drop in neutralisation titre against Omicron ranges from 2- to over 20-fold depending on the study and serum tested. This large range is not surprising, since we have shown in a large meta-analysis of the neutralisation of prior VOC that the drop in neutralisation against a variant varies dramatically between laboratories and assays, and also depends on the potency of the serum and limit of detection of the assay (3). Further, we identified that, after accounting for these laboratory and censoring effects, there were no significant differences between vaccines in the fold drop in neutralisation titres for each VOC. Thus, we found that to estimate neutralisation against a new variant it was most robust to aggregate a range of estimates from multiple laboratories rather than rely on a single laboratory result (3).

Collating data from the four studies listed above and accounting for censoring of neutralisation titres below the limit of detection (which is critical since many samples are below detection for Omicron) results in a combined estimate of the drop in neutralisation titre against Omicron of 9.7- fold (95% CI 5.5-17.1). This is shown in Figure 1A (grey line) along with the censored estimates of the fold drops by study (black lines) and within each cohort of a study (coloured lines).

**Figure 1.**
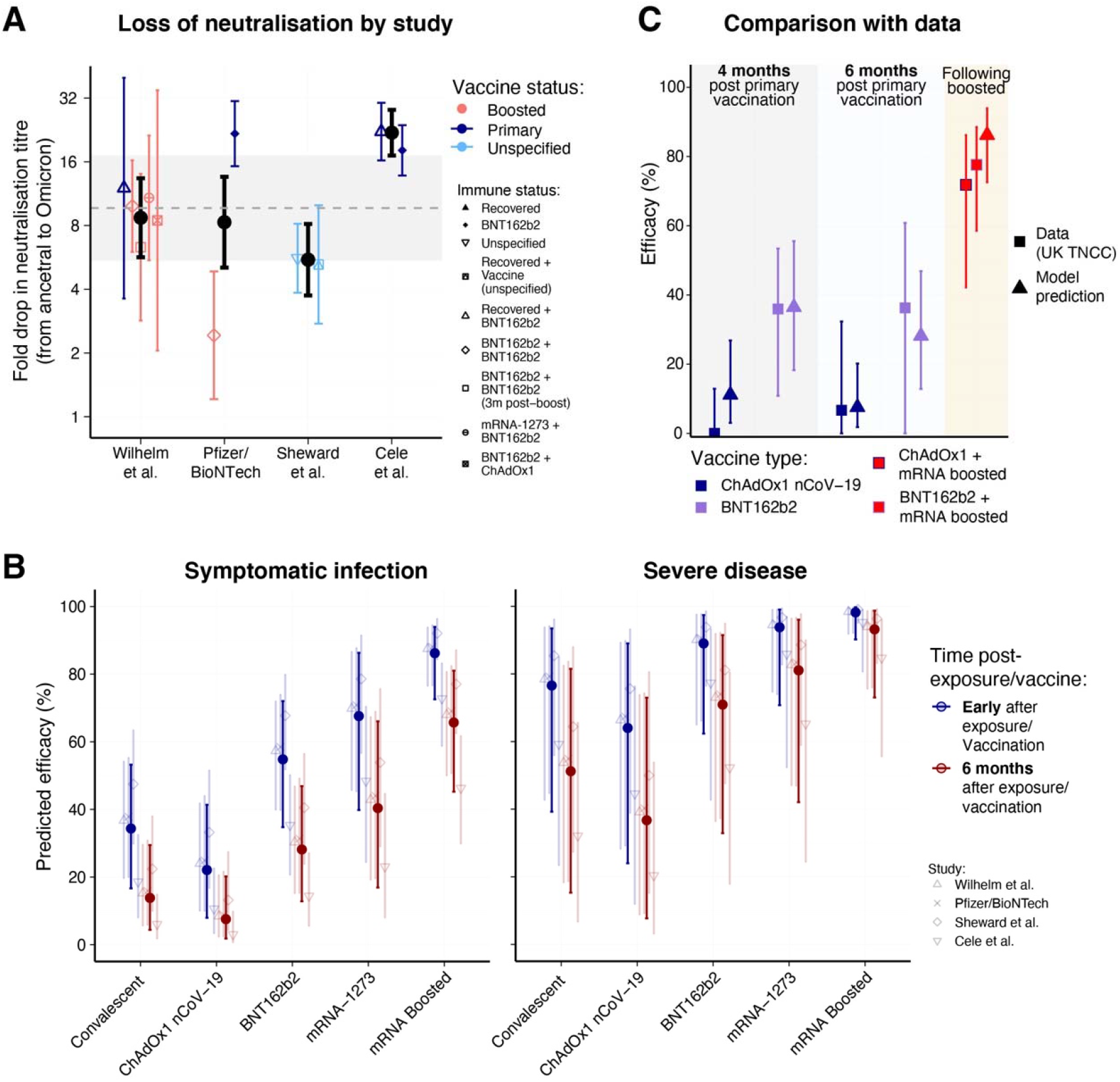
(A) Estimated fold drops in neutralisation titre against Omicron compared to the ancestral strain of SARS-CoV-2 across studies and cohorts available as of 13^th^ December 2021. Black dots indicate the geometric mean fold drop in neutralisation observed in each study across the different cohorts tested (with censoring at the limit of detection). All error bars indicate the 95% confidence intervals of the mean for each group. The grey dashed line indicates geometric mean of the studies (i.e. geometric mean of the black dots) and grey shading region indicates the 95% confidence interval of this mean of the studies. (B) Predictions of vaccine efficacy against Omicron for symptomatic (left) and severe (right) disease. Vaccine efficacies are shown early after exposure / vaccination (blue) and six months after exposure / vaccination (red). The neutralisation levels 6 months after infection, vaccination or boosting were calculated using a decay half-life of neutralising antibodies of 108 days (as reported in (3). mRNA boosted neutralisation levels are based on average levels reported in previously infected individuals boosted with an mRNA vaccine from (3). Error bars indicate 95% confidence intervals calculated by bootstrapping as in (3). (C) Comparison between estimates of Vaccine Efficacy against Omicron from a UK TNCC study (9) (squares) and our model estimate (triangles) at four months, six months and after boosting with an mRNA vaccine for ChAdOx1 nCoV-19 (blue) and BNT162b2 (purple).

Combining this estimate of the drop in neutralisation to Omicron (fig. 1A) we next estimate vaccine efficacy (or protection for convalescent individuals) for vaccines shortly after vaccination and six months later (2) (approach described in (3)). We estimate that six months after primary immunisation vaccine efficacy against Omicron has waned to 7.5% (95% CI: 1.8-20.2), 28.1% (95% CI: 12.8-46.9) and 40.4% (95% CI: 16.9-66) protection against symptomatic infection for ChAdOx1 nCoV-19, BNT162b2 and mRNA-1273 vaccines respectively and 36.7% (95% CI: 7.7- 73), 70.9% (95% CI: 32.9-91.5) and 81.1% (95% CI: 42.1-96) protection against severe symptoms for the same three vaccines (fig. 1B). A third booster dose with an existing mRNA vaccine (even though it targets the ancestral spike) has the potential to raise efficacy against Omicron to 86.2% (95% CI: 72.6-94) against symptomatic infection and 98.2% (95% CI: 90.2-99.7) against severe infection.

These estimates are in very good agreement with recent vaccine efficacy estimates from a Test Negative Case Control (TNCC) study published by the UK Health Security Agency (9). The comparison between the UK data and our model estimates is shown in (fig. 1C). Emerging data on the loss of neutralisation against the Omicron variant reveals considerable escape of neutralizing responses, but indicates that high levels of protection to symptomatic and severe infection is likely to be achieved by boosting with existing vaccines that target ancestral spike protein.

## Methods

### Estimating fold-drop in neutralisation titre for Omicron

Neutralisation titres from four available studies that compared titres against the ancestral and Omicron strain were either provided (4) or extracted from figures (5-8) using WebPlotDigitizer (10). This resulted in 175 measurements of neutralising antibody titres against the Omicron variant.

We calculated the geometric mean decrease in neutralisation titre in the different groups within each study and combined these to get an overall mean decrease for each study (comparing with ancestral virus). The overall mean was then calculated as the average of the study means (to avoid biasing studies that had more groups). We used censoring to account for neutralisation titres below the limits of detection of the assay, as described in (2) with limits of detection set at 1/25 (4), 1/45 (5) (limit not reported but inferred from (11) which is assumed to be the same assay) and 1/10 (6, 7).

### Predicting vaccine efficacy against Omicron

We have previously developed a model relating neutralisation levels to vaccine efficacy against symptomatic and severe SARS-CoV-2 infection for the ancestral strain of SARS-CoV-2. This was parameterised based on data from phase 1/2 and phase 3 trials (2). We have subsequently shown that this model is also predictive of vaccine efficacy against other VOC (3). We used this model to predict the efficacy of vaccines against Omicron using the average fold-change in neutralisation estimated above.

Briefly, vaccine efficacy for vaccine 𝒱 (*VE*_𝒱_) was estimated based on the (log_10_) mean neutralization titre as a fold of the mean convalescent titre reported for vaccine 𝒱 in phase 1/2 trials (*µ*_𝒱_), and the (log10) fold drop in neutralization titre to Omicron (*f*) using the equation:

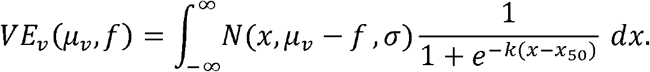

Here, *N* is the probability density function of a normal distribution with mean *µ* _𝒱_ -(*f)* and standard deviation σ, and *k* and *𝓍*_50_are the parameters of the logistic function relating neutralization to protection. All parameters excluding *f* were estimated previously as *σ* = 0.46, *k*= 3 and *𝓍*_50_*=* log_10_ 0.2 for symptomatic infection and 𝓍_50_= log_10_ 0.03 for severe and *µ* _𝒱_ -0,-0.27,0.38,0.62 and 1.08 and for early convalescent, ChAdOx1 nCoV-19, BNT162b2 and mRNA-1273 and boosted individuals respectively (2, 3). Note, the estimate of *µ* _𝒱_ for ChAdOx1 nCoV-19 is that associated with a 4-week interval between doses.

## Data Availability

All data produced in the present study are available upon reasonable request to the authors

## Funding and Ethics

This work was approved under the UNSW Sydney Human Research Ethics Committee (approval HC200242).

DSK, DC, and MPD are supported by NHMRC (Australia) Fellowship / Investigator grants. Some data in this study was supported by the Bill and Melinda Gates award INV-018944 (AS), National Institutes of Health award R01 AI138546 (AS), and South African Medical Research Council awards (AS). The funders had no role in study design, data collection and analysis, decision to publish, or preparation of the manuscript.

## Notes

### Competing Interest Statement

The authors have declared no competing interest.

### Author Declarations

This work was approved under the UNSW Sydney Human Research Ethics Committee (approval HC200242).

### Summary of Updates

Figures 1-3 combined. Vaccine specific efficacy estimates added into the text.

